# Establishing the operating conditions of ‘Ocula AI’ in capturing the pupil light reflex

**DOI:** 10.1101/2025.03.03.25323271

**Authors:** Sieu K. Khuu, Rebecca He, Brendan O’Brien

## Abstract

In recent years, rapid advances in technology have made it possible to monitor human health with personal handheld devices. While initially limited to measures of electrical skin potential for cardiac assessment, this has now expanded to include brain health by leveraging visual reflexes. In this study, we examine the effectiveness of a cutting-edge smartphone application, Ocula AI (Equinox), to capture and quantify the pupillary light reflex (PLR). In particular, the present study evaluates and compares the capability and operating range of Ocula AI, against an established clinical-standard device, the PLR-3000 pupillometer (NeurOptics). Both devices capture the PLR waveform providing estimates of key metrics such as such as latency, maximum and minimum pupils, and constriction and dilation velocities. The ability of Ocula AI to capture the PLR was assessed under different indoor illumination conditions (indicated by illuminance levels ranging from 0 to 1200 lux) in 16 healthy young adults. Our comparison and Bland-Altman analyses showed that Ocula AI effectively and reliably captured the PLR and estimated key features of the PLR waveform to a similar standard and with high agreement to the PLR-3000 device. Furthermore, Ocula AI was capable of capturing the PLR up to approximately 1000 lux, at which point the pupils are maximally constricted. These results provided preliminary evidence of the utility and potential benefit of mobile devices in providing accurate and easy estimation of the PLR.

## Introduction

The pupillary light reflex (PLR) is a rapid and automatic response of the pupil of the eye to changes in light intensity. The pupils constrict (miosis) to the onset of bright light, but dilate (mydriasis) under low light levels. This reflex is crucial to vision as it regulates the amount of light entering the eyes to ensure optimal vision and protects the retina from excessive light exposure (see Belliveau et al., 2025).

The PLR reflects the function of the autonomic nervous system, which regulates the antagonistic actions of the iris sphincter (miosis – parasympathetic) and the dilator (mydriasis – sympathetic) muscles of the eye to control the amount of light entering the eye. Though this action is simple, it is underpinned by a complex array of pathways and neural networks, brainstem, and spinal cord (see Bergamin & Kardon, 2003; Yoo & Mihaila, 2025). The PLR is mediated by the brain’s afferent (sensory; visual) and efferent (motor; miosis-mydriasis) pathways, forming a reflex arc. The afferent pathway conveys sensory information regarding the light intensity, particularly signals from intrinsic photosensitive retinal ganglion cells(ipRGC) in the eye to the brain. In contrast, the efferent pathway conveys motor commands from the brain to the muscles controlling the pupil, particularly the iris sphincter and dilator muscles, which control pupil constriction and dilation respectively. Previous studies have extensively reported abnormalities in the PLR from normal ageing (e.g. Belliveau et al., 2025), in conditions such as Parkinson’s and Alzheimer’s disease (e.g. Sparks et al., 2023; Tsitsi et al., 2023), and disorders such as chronic fatigue, COVID-19 infection, and traumatic brain injury (TBI), particularly concussion (see e.g. Belliveau et al., 2025; Boulter et al., 2021; Ciuffreda et al., 2017; El Ahmadieh et al., 2021). Such studies also highlight the simplicity of measuring the PLR and its utility as a biological marker for brain-related diseases and disorders.

The PLR is typically assessed in the clinical setting using the relative afferent pupillary defect test (RAPD). Here, light is presented in sequence to each eye, and the pupil size of both eyes is compared. However, this test is highly subjective and dependent on the skill and experience of the clinician. Further, it does not capture key temporal characteristics of pupil size changes from light exposure (see Figure 1). Typically, in response to exposure to a bright light source, the pupil quickly constricts (after a brief delay or latency) from a baseline pupil size (maximum pupil size) to reach a maximum constriction size (minimum pupil size). After the light source is turned off, the pupil typically slowly dilates back to baseline. The utility of measuring the PLR waveform is that one or a combination of these temporal aspects is known to be affected by brain-related changes caused by disease, injury and neurotoxicity due to exposure to drugs or chemicals. For example, concussion is known to lead to slower PLR latencies and longer constriction and dilation velocities (see Carrick et al., 2021 for a review), and this has also been reported in patients with Parkinson’s disease (e.g., Mosaad, Hovis & Almeida, 2022). We have also shown alterations in latency and maximum constriction size from pesticide exposure (Jiménez-Barbosa et al., 2024), revealing the utility of objectively measuring aspects of the PLR waveform.

**Figure 1.**
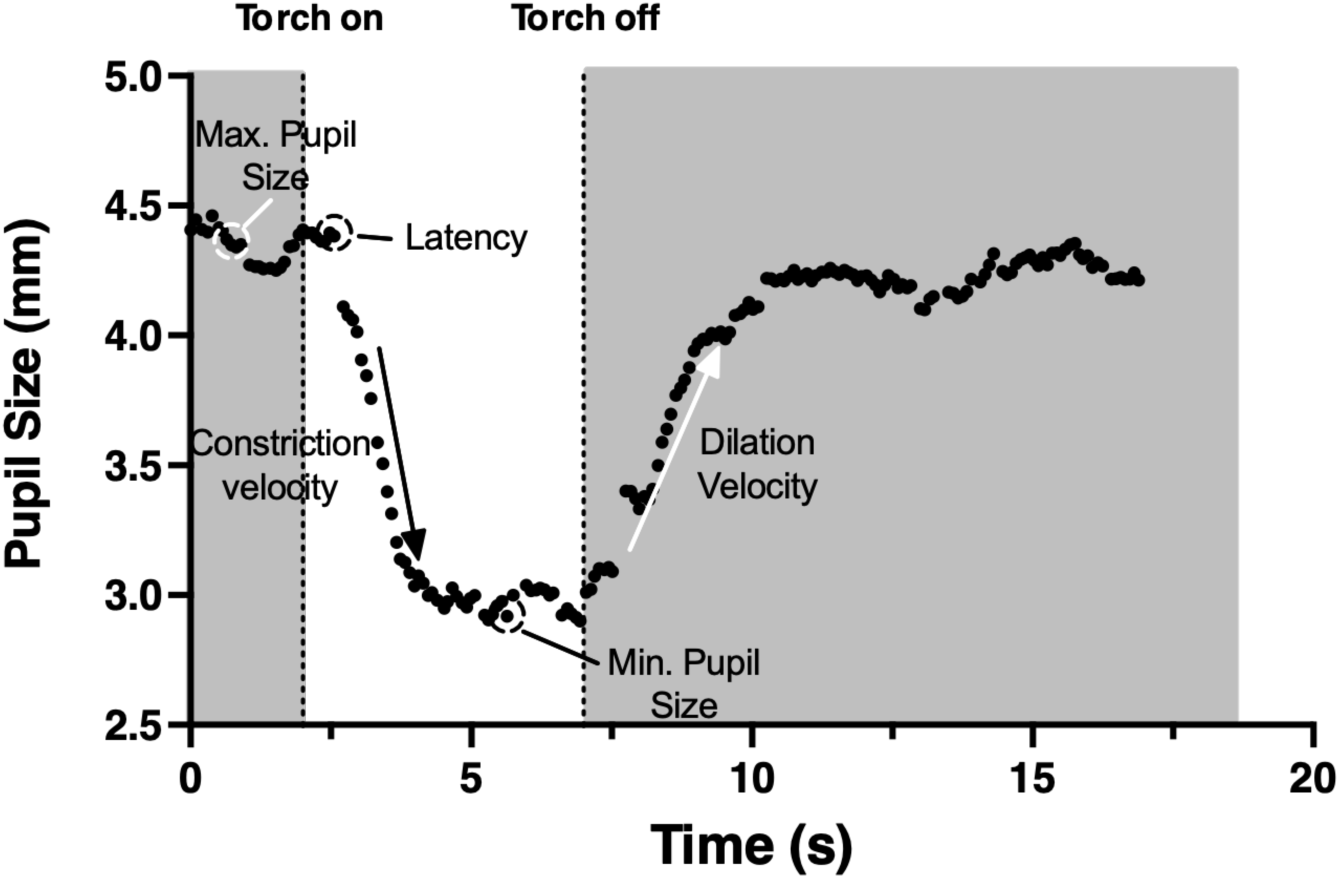
The PLR waveform with key features of the waveform is highlighted. Prior to the onset of the torch, the pupil remains at its maximum pupil size. With the onset of the torch/light, and after a brief delay/latency, the pupil rapidly constricts at a particular velocity to reach a minimum pupil size. After the torch/light is turned off, the pupil gradually dilates at a particular velocity (dilation velocity). After which the pupil remains at a stable/constant size.

Dedicated clinical devices capable of capturing the PLR waveform exist, e.g., the PLR-3000 device (by NeurOptics), however, such technologies are expensive, not easy to use, and not widely available. This device also utilises older technologies and infrared lighting which may lead to low resolution images and image blur which may affect recording accuracy (Dulski et al., 2010). Recognising the need for more assessable, affordable, and easier-to-use technologies to measure the PLR, mobile device “apps” are now available on both Android and Apple platforms. Such apps capitalise on the capability of mobile devices with camera technologies that employ the devices ‘flash’/torch to generate a PLR and capture high-resolution video recordings of the PLR. Further, proprietary software available in these apps can analyse the PLR waveform to estimate the temporal aspects of the PLR on demand. However, at present, the use of such apps is limited by thorough research regarding their accuracy (relative to a gold standard), operating capabilities, particularly across a range of conditions, and most critically, the ambient light of the testing environment and the accuracy in estimating the PLR waveform. Additionally, at present, device specific PLR recording apps lack normative comparison data that considers the PLR’s dependency on factors such as eye colour, age, and gender.

The Ocula AI app by Equinox Technologies ™ is a mobile app designed to utilise the nascent technologies available on newer devices to provide more accurate estimates of the PLR, and future diagnostic capabilities through artificial intelligence models to detect abnormalities in the PLR. Particularly, the app utilises the increasingly powerful onboard processing units of modern mobile devices and high-resolution cameras to record the pupil and estimate parameters of the PLR, which may provide more accurate PLR measurements over older app technologies. Different settings allow for customised measurements of the PLR which take into consideration pupil colour, sex, and age. As this is a new technology with good potential, in the present study, we tested Ocula AI’s capability of capturing the PLR and compared its outputs to the PLR-3000 device, which is a clinical standard. In particular, we sought to establish the minimum and maximum lighting conditions under which Ocula AI operates and the app’s accuracy in estimating the PLR.

## Methods

### Participants

Sixteen normal participants who met the study’s inclusion criteria were recruited for the study. The mean age of participants was 27 years (range: 20 – 37), and 10 had brown eyes, 2 had green eyes, and 4 had blue eyes. Participants were drawn from the local University of New South Wales (UNSW) population. All had normal or corrected to normal visual acuity, were of good general health, and had no self-reported history of traumatic brain injury or any neurocognitive disorders such as Parkinson’s and Alzheimer’s disease. Participants with pupillary abnormalities such as Adie’s Tonic pupil and Anisocoria were excluded from the study, as well as individuals with self-reported light-induced migraines or photophobia. Human research ethics was obtained and approved by the UNSW ethics committee, and the research conformed to the Declaration of Helsinki on human research.

## Procedure

This was a laboratory-based study in which the PLR was measured in normal participants for different light levels in a light-controlled room. Interested participants were advised to provide written consent. Once consent was obtained, participants underwent the following assessments/tests to ensure they met the selection criteria. 1) Best visual acuity at a distance and near was recorded using handheld vision charts (Test Chart 2000 Pro). 2) Screening for relative afferent pupillary defect (RAPD) using the swinging light test. 3) Colour Vision test using Ishihara colour plates. 4) Extra-ocular motility (Broad H Test). 5) Cover test. Participants were required to pass all tests.

### PLR testing using the Ocula AI app and PLR-3000

Eligible participants proceeded to have their PLR measured using the Ocula AI app installed on an Apple iPhone 14. The setup and use of Ocula AI is straightforward – through an intuitive graphics user interface, the app allows for customized testing that includes control over the duration of the light flash and measurement. Additionally, the app registered the following information: Age, Sex, Ethnicity, and Eye Colour as these factors are known to influence and must be considered when interpreting the data (Sharma et al., 2016).

The participant was seated, and his/her head stabilised by a head and chin rest. The device containing the Ocula AI app was positioned 20 cm from the participant at eye level and held steady using a tripod and clamp. The participant fixated on the torch, and Ocula AI proceeded to record the pupil simultaneously for both left and right eyes over a 17-second period using the following steps (see Figure 1): At the onset of recording, Ocula AI recorded the pupil for 2 seconds without the torch on. Then, a bright light source (device torch – 200 cd/m^2^) was turned on and presented for 5 seconds to induce a PLR and then extinguished. The recording continued for 10 seconds to capture the recovery of the pupil.

Ocula AI is a smartphone app that captures video of the subject’s eyes at 60 frames per second (fps) with the onboard camera and processes it with a convolutional neural network model. Absolute pupil dimensions are calibrated by comparing them with iris diameter, which is nearly constant within the human population (e.g. Belliveau et al., 2025). Calibration was done per eye on a frame-by-frame basis to minimize measurement error due to the physical instability of the user or the subject during recording. The data were smoothed and artefacts (e.g. eye blinks) were removed using proprietary algorithms. After measurement, estimates of the maximum pupil size (prior to the onset of constriction), latency, and constriction and dilation velocities were immediately derived from the PLR waveform using proprietary algorithms and displayed on a summary screen (see Figure 2B). First and second derivatives of the data are calculated to measure the velocity and acceleration of pupil constriction/dilation. Latency is calculated using the approach outlined in Bergamin & Kardon (2003). High resolution video recordings of the eye and PLR response were also stored on the device as a source for future re-analysis if needed.

**Figure 2.**
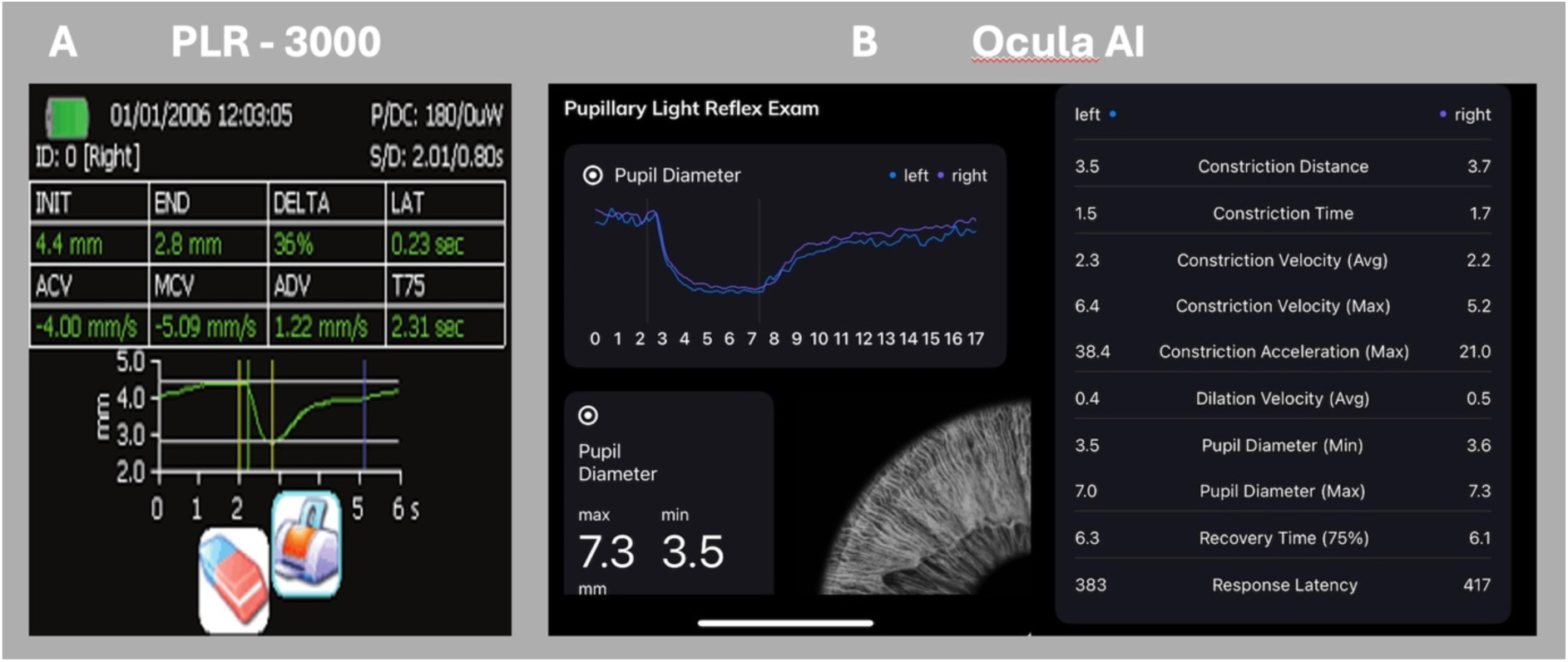
Output and interface for the PLR-3000 (A) device and Ocula AI. Both devices provide immediate estimation of PLR metrics (e.g., maximum and minimum pupil diameters, latency and velocity) which are reported after testing.

The steps above were repeated to measure the PLR for eight room illumination levels of 0, 50, 100, 200, 400, 600, 800, 1000, and 1200 lux in a randomised order. This light range spans the typical light levels of an indoor clinical or office space (see Brown et al., 2022). The light source was a 24W LED lamp set to a colour temperature of 5000K corresponding to daylight, with adjustable intensity. The lamp was positioned directly in front so that it fully illuminated the participant’s face. A Minolta lux meter (T-10MA) was fixed adjacent to and level with the participant’s eye facing the light source to provide a close indication of the illumination intensity entering the eyes. Breaks of at least 1 minute, or longer if the participant requested it, were provided between measurements to ensure that participants remained comfortable.

Additionally, the PLR was measured using the PLR-3000 device for both left and right eyes under the same test settings as Ocula AI. The eye tested first was randomised from trial to trial. This device takes infrared recordings of the PLR and onboard propriety software (see Armas et al., 2024; McKay et al., 2020 for a review) provides estimates of the same aspects of the PLR as Ocula AI allowing for comparison. Outputs are displayed on the device screen (see Figure 2A) but can also be downloaded as a CSV file via Bluetooth. Note that this device measures the PLR using a closed eyepiece to ensure dark conditions, and therefore, testing under different lighting conditions is not possible.

As mentioned, the Ocula AI and PLR-3000 devices provided estimates of the maximum and minimum pupil size, latency (i.e., the beginning of pupil constriction), and constriction and dilation velocities from the PLR waveform, which can be read from the devices or downloaded via Bluetooth. These measures were extracted from the devices and recorded in a spreadsheet for statistical analysis. Data were analysed using the GraphPad Prism (version 11) software. The Kolmogorov-Smirnov test was used to assess for normality and t-tests were used to infer statistical significance (α = 0.05).

## Results

All data passed tests of normality and there was no statistical difference (t-test) between left and right eyes across all PLR measurements for both Ocula AI and the PLR-3000 device and accordingly, they were combined. The ROUT method (see Motulsky & Brown, 2006) was used to identify outliers (Q-quotient of 1 %). Data from one participant was excluded from the analysis as they withdrew during testing.

### Maximum and Minimum Pupil Size

Results from the left and right eyes were pooled for both devices as there was no statistically significant difference in PLR waveforms. In Figure 3A, average, maximum, and minimum pupil size (diameter (in mm) different coloured symbols) obtained from PLR measurements are plotted as a function of room illuminance (in lux). Both metrics are plotted to indicate how the pupils changed from baseline (maximum pupil size) to maximum constriction (minimum pupil size) due to light exposure. Best-fit exponential decay functions are shown as solid lines (average R^2^=0.88). Also shown in Figure 3A are the average maximum and minimum pupil sizes (dashed horizontal) obtained using the PLR-3000 device. As mentioned, this device uses a closed aperture and operates with no background illumination (equivalent to 0 lux), hence there was only one measure.

**Figure 3.**
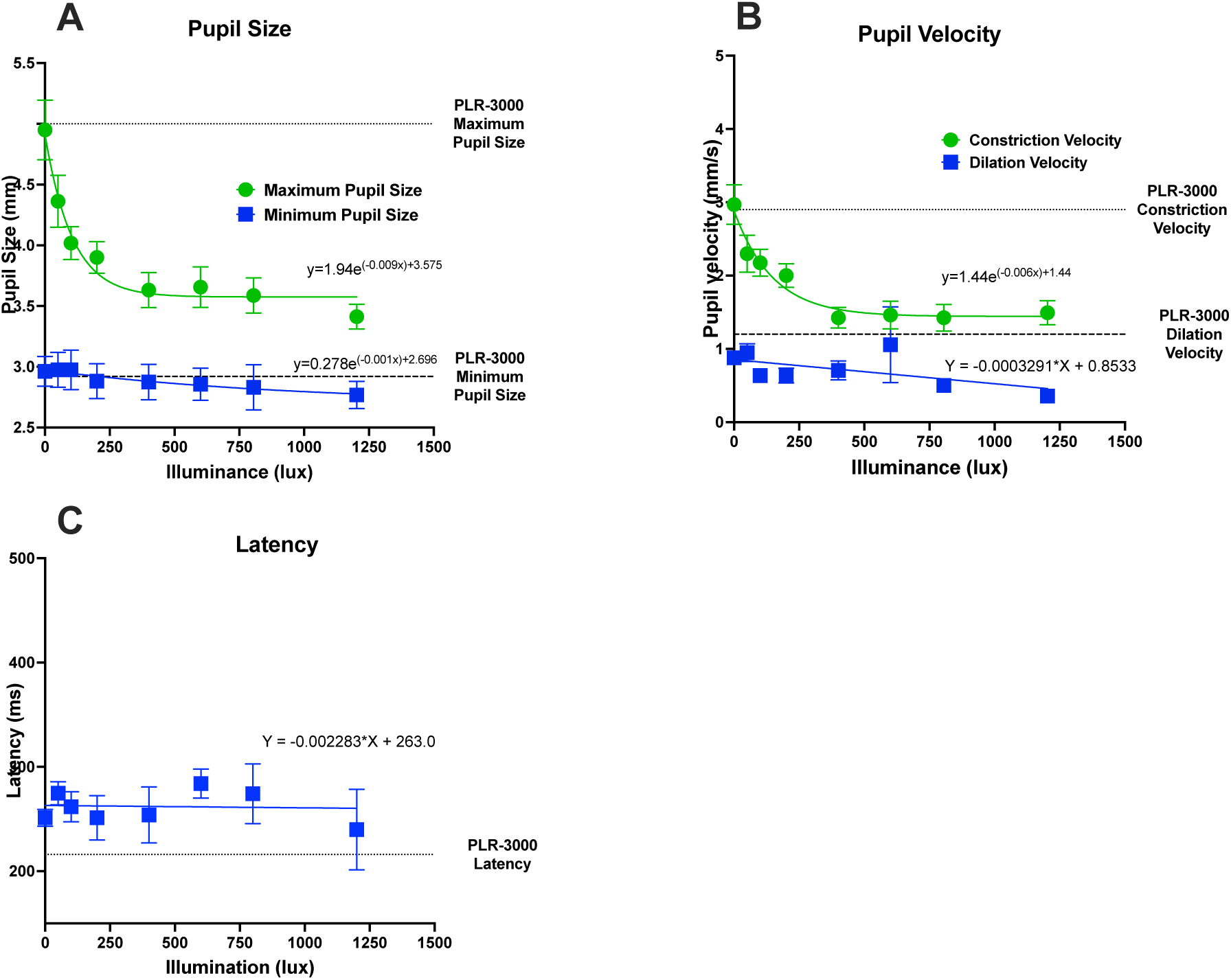
Average, maximum and minimum Pupil Sizes (A) and Velocities (B), and latency (C) obtained using Ocula AI are plotted as a function of room illuminance (lux). Error bars signify one standard error of the mean. Average measurements obtained from the PLR-3000 device are shown as dashed horizontal lines. The exponential/line of best fits is shown as solid lines.

Figure 3A shows that at the lowest light levels (0 lux), maximum and minimum pupil sizes were comparable to the PLR-3000 device. As expected, for measurements obtained using Ocula AI, increasing room illuminance reduced maximum pupil size (following an exponential decay), as the pupils are naturally constricted due to higher light energy in the environment. This finding has been well documented in the literature (e.g., see Hu, Hisakata & Kaneko, 2020) and naturally reflects the physiological regulation of light entering the eyes dependent on room illuminance. The minimum pupil size (due to pupil constriction in response to the flash of light) remained unchanged over the room illuminance range measured in the present study and is indicative of a physiological constriction limit. Our findings showed that Ocula AI effectively captured expected changes in pupil sizes due to a PLR over a broad range of room illumination.

Statistical comparison (paired t-test) between the PLR-3000 device and Ocula AI data obtained at an illuminance of 0 lux were performed separately for maximum and minimum pupil size conditions. As mentioned, the PLR-3000 device operates with a closed eyepiece and thus, operates with no background illuminance, and this is equivalent to Ocula AI testing at a background illuminance of 0 lux. These pairwise analyses showed no significant difference for both maximum pupil size, t(15)=0.897 p=0.383 and minimum pupil size: t(15)=0.032, p=0.974 conditions. These results indicate that both devices are providing similar estimates of pupil size at an illuminance level of 0 lux.

Further, Bland-Altman plots (Riffenburgh & Gillen, 2020) of these data was constructed by plotting the difference between Ocula AI and the PLR 3000 device as a function of the average of both devices (see Figures 4A and B). This plot provides visual confirmation of the high agreement between both devices with most data points within the 95 % agreement bands (dashed lines) and minimal bias (Maximum pupil size: -0.08 mm, SD:0.390; Minimum pupil size: 0.003, SD=0.38).

**Figure 4:**
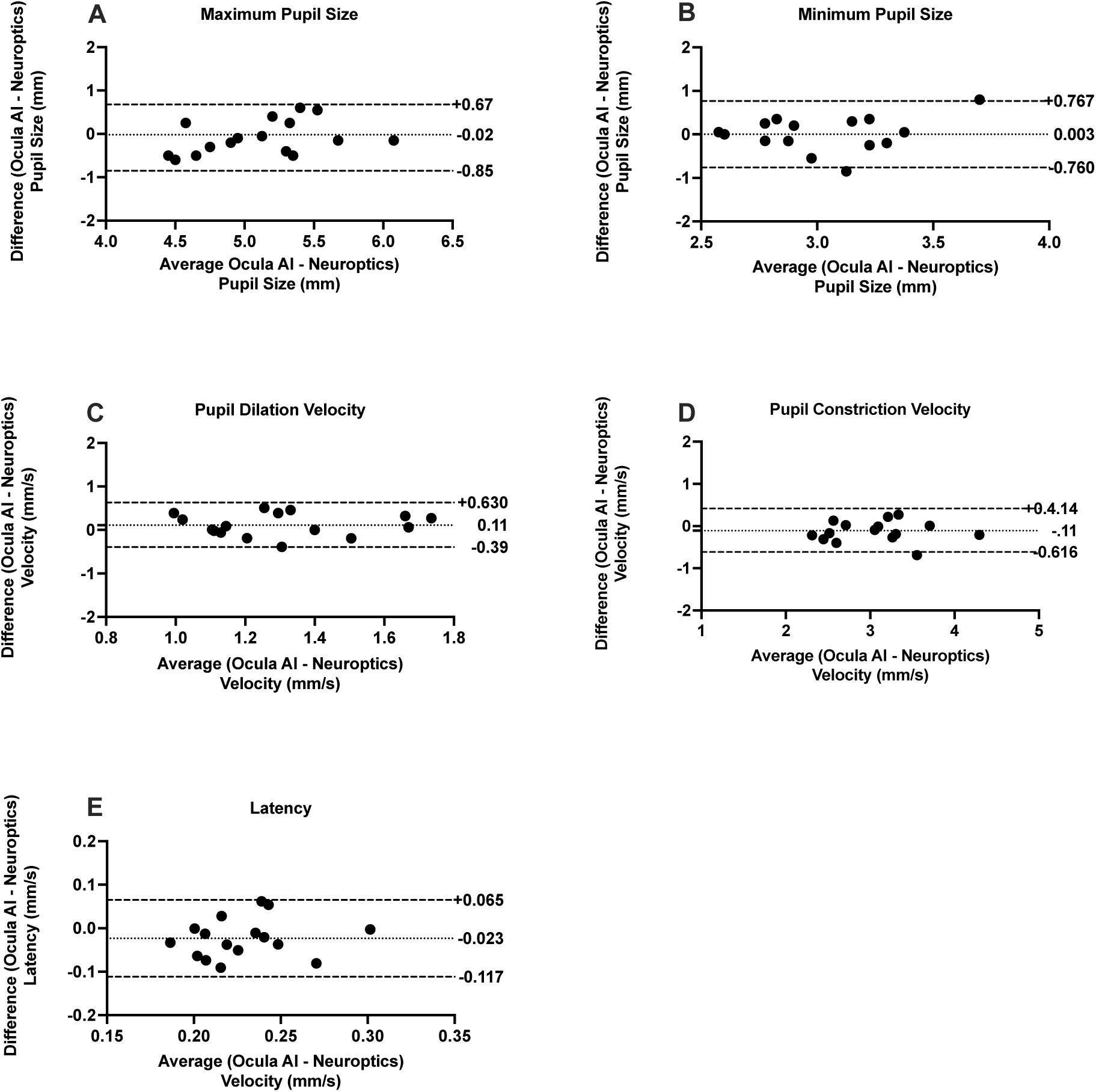
Bland Altman plots comparing the difference between Ocula AI and the PLR-3000 device for PLR metrics of maximum and minimum Pupil Sizes (A & B), Pupil dilation and constriction velocities (C & D), and Latency (E). Dotted lines is indicative of the average bias, and dashed lines are indicative of upper and lower 95 % confidence agreement bands.

### Constriction and Dilation Velocities

In Figure 3B, mean absolute constriction and dilation velocities (mm/s) of the PLR are plotted as different symbols as a function of room illuminance. Equivalent values obtained from the PLR-3000 device are indicated by dashed horizontal lines. Change in velocities as a function of room illumination was approximated by an exponential decay function, but this trend was evident only for constriction velocity values. Increasing illuminance reduced the constriction velocity, perhaps because baseline pupils are smaller (see Figure 3A) due to the increased background illumination. In contrast, dilation values remained unchanged as indicated by a linear fit in which the slope (-0.001) was not significantly different from 0 (F(1,110)=3.207, p=0.0761). This finding indicates that PLR recovery (after the offset of the flash of light) may not be dependent on background illumination.

At equivalent lowest illuminance levels (0 lux), velocity values obtained from Ocula AI were similar and not statistically different (Constriction Velocity: t(15)=1.272, p=0.223, Dilation Velocity: t(15)=1.680, p=0.114) to measurements obtained with the PLR-3000 device. Bland-Altman plots (See Figure 4C & D) showed high agreement between both devices with most values within the 95 % agreement confidence intervals, and little bias (Constriction Velocity: -0.100 mm/s, SD:0.263; Dilation Velocity: 0.118mm/s, SD:0.261).

### Latency

Average latency values obtained using Ocula AI are plotted as a function of room illuminance in Figure 3C. As shown in Figure 3C, latency values recorded by Ocula AI did not systematically change with room illuminance, indicating that the time taken for a pupillary contraction is not dependent on the level or room illuminance. A linear fit to these data showed that the slope of the best fitting line (see solid line in Figure 3C) did not significantly differ from 0 (F(1,96)=0.015, p=0.903). Note that latency values obtained from Ocula AI are on average approximately 20 ms longer than those obtained from the PLR-3000 device. This difference is observed across all illuminance values and may reflect a difference in methods used to estimate latency between the two technologies. However, a paired t-test (between Ocula AI measurements at 0 lux and the PLR-3000 device) showed that this difference was not significant (t(15)=1.897, p=0.077). Bland-Altman plots of the difference in latency between the devices show this small bias: -0.020, SD: 0.042, however, there remains a high agreement between both devices (see Figure 4E).

## Discussion

In the present study we evaluated and compared PLR measurements between Ocula AI, a smartphone app, and the PLR-3000 device, a dedicated device designed for clinical measurements of the PLR. We find that under similar operating conditions, the outputs of both devices are similar with good agreement. However, Ocula AI can operate under a range of lighting conditions spanning indoor conditions (100 – 1000 lux units) and is effective in quantifying the PLR over a broad range of illuminances. The reported good agreement between the Ocula AI and the PLR-3000 device may be representative of improvements in mobile app software design and smartphone hardware compared to older systems and technologies (McKay et al. 2020).

The comparable performance of Ocula AI, in addition to its ease of use, portability, and cost effectiveness, presents itself as an effective way of quantifying the PLR outside of the clinical setting as a means of detecting brain-related change and recovery. The utility of this approach lies in the well-established relationship between changes in the PLR and brain-related injury such as traumatic brain injury. Particularly, concussion/mild traumatic brain injury associated with activities such as contact sports, falls, and blasts can be debilitating. Quick detection of signs of concussion (such as abnormalities in the PLR) may allow medical and support staff to provide immediate assistance or intervention to prevent further injury and inform decisions regarding continuing an activity. Additionally, a period of rest (typically 1 – 2 weeks) is mandatory before a return to work or play. However, recovery over this period is typically not monitored running the risk of a return to work/play before full recovery is achieved. Importantly, technologies such as Ocula AI provide an effective way of detecting signs of concussion through abnormal PLR, and a means of monitoring recovery (particularly at home) without the need for medical assessment by clinical devices and visits.

Though promising as a portable and reliable means of estimating the PLR, some issues must be considered regarding the use of mobile apps. Note that in the present study, PLR measurements were obtained under controlled laboratory conditions, but on-field testing may introduce factors that can impact measurement. First, as shown in the present study and well documented in the literature, the lighting conditions are critical for PLR measurements. Particularly, given the open environment in which smartphone apps measure the PLR, testing needs to be performed in low light levels (typically indoors), and not under high bright illuminance levels that are inherent to the outdoors. This is not a limitation of mobile devices, but rather a physiological limit of the visual system as the pupils are already maximally constricted at high illuminance levels. Secondly, the device is handheld and requires the operator to maintain a steady pose over the measurement period to ensure that the eyes and pupils are recorded. Excessive movement may affect the quality and reliability of the recording. Third, while this study establishes the capabilities of a smartphone app in quantifying the PLR, there is a need for further validation and testing (under a broad range of environmental conditions) to derive normative data, particularly across a range of illuminances. These data, in addition to thoroughly establishing the real-world operating characteristics for its effective use, are necessary if such technologies are intended for diagnostics purposes and clinical use.

The performance of Ocula AI has not been systematically investigated in a clinical population. Further study in a clinical population is recommended to further establish the utility of Ocula AI as a detection/diagnostic tool for injuries or diseases that disrupt normal brain function. Further, comprehensive measurement in larger and more diverse cohorts of participants is needed to establish norms for clinical comparison and the development of AI technologies that support the detection/diagnosis of brain-altering maladies.

## Data Availability

All data produced in the present study are available upon reasonable request to the authors

## References

Armas, S., Jehu, D., Bolgla, L., & Dutton, F. (2024). Examining the inter -rater and inter-trial reliability of the NeuroOptics -3000 pupilometer to measure pupillary light response parameters. Archives of Physical Medicine and Rehabilitation, 105(4), e184. 10.1016/j.apmr.2024.02.636

Belliveau, A. P., Somani, A. N., & Dossani, R. H. (2025). Pupillary Light Reflex. In StatPearls. StatPearls Publishing. http://www.ncbi.nlm.nih.gov/books/NBK537180/

Bergamin, O., & Kardon, R. H. (2003). Latency of the Pupil Light Reflex: Sample Rate, Stimulus Intensity, and Variation in Normal Subjects. Investigative Ophthalmology & Visual Science, 44(4), 1546–1554. 10.1167/iovs.02-0468

Boulter, J. H., Shields, M. M., Meister, M. R., Murtha, G., Curry, B. P., & Dengler, B. A. (2021). The Expanding Role of Quantitative Pupillometry in the Evaluation and Management of Traumatic Brain Injury. Frontiers in Neurology, 12, 685313. 10.3389/fneur.2021.685313

Brown, T. M., Brainard, G. C., Cajochen, C., Czeisler, C. A., Hanifin, J. P., Lockley, S. W., Lucas, R. J., Münch, M., O’Hagan, J. B., Peirson, S. N., Price, L. L. A., Roenneberg, T., Schlangen, L. J. M., Skene, D. J., Spitschan, M., Vetter, C., Zee, P. C., & Wright, K. P. (2022). Recommendations for daytime, evening, and nighttime indoor light exposure to best support physiology, sleep, and wakefulness in healthy adults. PLoS Biology, 20(3), e3001571. 10.1371/journal.pbio.3001571

Carrick, F. R., Azzolino, S. F., Hunfalvay, M., Pagnacco, G., Oggero, E., D’Arcy, R. C. N., Abdulrahman, M., & Sugaya, K. (2021). The Pupillary Light Reflex as a Biomarker of Concussion. Life, 11(10), Article 10. 10.3390/life11101104

Ciuffreda, K. J., Joshi, N. R., & Truong, J. Q. (2017). Understanding the effects of mild traumatic brain injury on the pupillary light reflex. Concussion, 2(3), CNC36. 10.2217/cnc-2016-0029

Dulski, R., Powalisz, P., Kastek, M., & Trzaskawka, P. (2010). Enhancing image quality produced by IR cameras. Electro-Optical and Infrared Systems: Technology and Applications VII, 7834, 283–291. 10.1117/12.864979

El Ahmadieh, T. Y., Bedros, N., Stutzman, S. E., Nyancho, D., Venkatachalam, A. M., MacAllister, M., Ban, V. S., Dahdaleh, N. S., Aiyagari, V., Figueroa, S., White, J. A., Batjer, H. H., Bagley, C. A., Olson, D. M., & Aoun, S. G. (2021). Automated Pupillometry as a Triage and Assessment Tool in Patients with Traumatic Brain Injury. World Neurosurgery, 145, e163–e169. 10.1016/j.wneu.2020.09.152

Hu, X., Hisakata, R., & Kaneko, H. (2020). Effects of stimulus size, eccentricity, luminance, and attention on pupillary light response examined by concentric stimulus. Vision Research, 170, 35–45. 10.1016/j.visres.2020.03.008

Jiménez-Barbosa, I. A., Grajales Herrera, D., Rodríguez Alvarez, M.F., & Khuu, S. K. (2024). Pupil size change in agricultural workers exposed to pesticides. Clinical and Experimental Optometry, 107(8), 795–800. 10.1080/08164622.2023.2294810

Mosaad, A, Hovis, J. K., Almeida Q.J. (2022). Pupil light reflex in Parkinson’s disease patients with and without freezing of gait symptoms. Saudi Journal of Ophthalmology 35(4), 332–340. 10.4103/1319-4534.347306

McKay, R. E., Kohn, M. A., Schwartz, E. S., & Larson, M. D. (2020). Evaluation of two portable pupillometers to assess clinical utility. Concussion, 5(4), CNC82. 10.2217/cnc-2020-0016

Motulsky, H. J., & Brown, R. E. (2006). Detecting outliers when fitting data with nonlinear regression—A new method based on robust nonlinear regression and the false discovery rate. BMC Bioinformatics, 7, 123. 10.1186/1471-2105-7-123

Riffenburgh, R. H., & Gillen, D. L. (2020). 27—Techniques to Aid Analysis. In R.H. Riffenburgh & D. L. Gillen (Eds.), Statistics in Medicine (Fourth Edition) (pp. 631– 649). Academic Press. 10.1016/B978-0-12-815328-4.00027-9

Sharma, S., Baskaran, M., Rukmini, A. V., Nongpiur, M. E., Htoon, H., Cheng, C.-Y., Perera, S. A., Gooley, J. J., Aung, T., & Milea, D. (2016). Factors influencing the pupillary light reflex in healthy individuals. Graefe’s Archive for Clinical and Experimental Ophthalmology, 254(7), 1353–1359. 10.1007/s00417-016-3311-4

Sparks, S., Pinto, J., Hayes, G., Spitschan, M., & Bulte, D. P. (2023). The impact of Alzheimer’s disease risk factors on the pupillary light response. Frontiers in Neuroscience, 17, 1248640. 10.3389/fnins.2023.1248640

Tsitsi, P., Nilsson, M., Waldthaler, J., Öqvist Seimyr, G., Larsson, O., Svenningsson, P., & Markaki, I. (2023). Pupil light reflex dynamics in Parkinson’s disease. Frontiers in Integrative Neuroscience, 17, 1249554. 10.3389/fnint.2023.1249554

Yoo, H., & Mihaila, D. M. (2025). Neuroanatomy, Pupillary Light Reflexes and Pathway. In StatPearls. StatPearls Publishing. http://www.ncbi.nlm.nih.gov/books/NBK553169/

